# Linking cortical lesions to metabolic changes in multiple sclerosis using 7T proton MR spectroscopy

**DOI:** 10.1101/2023.08.18.23294260

**Authors:** Mads Alexander Just Madsen, Michal Považan, Vanessa Wiggermann, Henrik Lundell, Morten Blinkenberg, Jeppe Romme Christensen, Finn Sellebjerg, Hartwig Roman Siebner

## Abstract

**Importance:** Cortical lesions contribute to disability in multiple sclerosis (MS) but their impact on regional neurotransmitter levels remains to be clarified.

**Objective:** To test the hypothesis that cortical lesions in MS alter the regional concentrations of the main excitatory and inhibitory neurotransmitters, glutamate and gamma-aminobutyric acid (GABA), in the affected cortex.

**Design:** Prospective, cross-sectional, observational proton MR-spectroscopy (^1^H-MRS) and structural MRI study at 7T.

**Setting:** Data were collected at a single center between August 2018 and September 2020.

**Participants:** A volunteer sample of 57 MS patients and 38 healthy participants were screened for participation in the study. 50 MS patients and 28 healthy participants were included. In the final cohort, three patients and five healthy participants were excluded due to drop out (n=6) or insufficient data-quality (n=2).

**Exposures:** Two-voxel 7T ^1^H-MRS covering the right and left sensorimotor hand areas (SM1-HAND) and high-resolution structural brain 7T MRI.

**Main outcome:** Regional concentrations of glutamate and GABA in SM1-HAND and their relation to cortical lesion volume within the MRS voxel.

**Results:** Data from 34 relapsing remitting (RR) and 13 secondary progressive (SP)MS patients (mean +/− standard deviation, 45.1 +/− 12.5 years, 31 female) along with 23 age- and sex-matched healthy participants (44.4 +/− 13 years, 15 female) entered data-analyses. Patient data were pooled to assess the relationship between cortical lesion volume and neurotransmitter levels. Larger cortical lesion volume within SM1-HAND was associated with higher regional glutamate (0.61 +/− 0.21 log(mm^3^), P=0.005) and lower regional GABA (−0.71 +/− 0.27 log(mm^3^), P=0.01) concentration. Between-group comparison showed that glutamate concentration within the SM1-voxel was reduced in SPMS patients compared to healthy participants (−0.75 +/− 0.24 mM, P=0.004) and RRMS patients (−0.55 +/− 0.22 mM, P=0.04), while regional GABA levels did not differ among groups.

**Conclusion:** Our results link cortical lesion load in SM1-HAND with regional glutamate and GABA levels in patients with RRMS and SPMS, showing a shift in balance between regional excitatory and inhibitory neurotransmitters towards increased excitation with increasing cortical lesion volume. Between-group comparisons provide preliminary evidence that a progressive disease course may be associated with a decrease in cortical glutamate levels.

**Key points:** **Question:** How do cortical lesions change the regional metabolic profile in multiple sclerosis?

**Findings:** This observational cross-sectional study employed voxel-based proton MR-spectroscopy (^1^H-MRS) of the primary sensorimotor hand areas (SM1-HAND) at ultra-high field (7T) to show that cortical lesions alter regional concentrations of glutamate and gamma-aminobutyric acid (GABA) in patients with multiple sclerosis. We found that higher regional glutamate concentrations were associated with larger regional cortical lesion volume, whereas higher GABA concentrations were associated with lower regional cortical lesion volume.

**Meaning:** These findings suggest that cortical lesions shift the regional excitation-inhibition balance towards excitation.

## Introduction

Multiple sclerosis (MS) is a chronic neuroinflammatory disease of the central nervous system.^1^ Focal white matter demyelination results in disseminated lesions that can be visualized with standard clinical MRI examinations.^2^ MS is not limited to focal white-matter lesions but also includes diffuse damage of radiologically normal appearing tissue and focal lesions in grey matter.^3,4^ Cortical lesions have become an important clinical marker of both physical^5,6^ and cognitive^5,7^ impairment in MS. Structural brain MRI at ultra-high field strength (7T) greatly increases the sensitivity of MRI to detect cortical lesions.^8–10^ Employing high-resolution structural 7T MRI, we recently showed that the presence of cortical lesions in the sensorimotor hand area (SM1-HAND) makes a specific contribution to sensorimotor disability in the contralateral hand.^11^ Determining the pathophysiology of cortical lesions is thus an important step towards the development of novel therapies targeting cortical involvement in MS.

Proton MR spectroscopy (^1^H-MRS) can quantify the regional concentrations of brain metabolites related to regional MS pathology revealed by structural MRI. A prominent peak in the ^1^H-MRS spectra reflects the regional concentration of N-acetylaspartate (NAA). NAA is a marker of neuronal density and viability^12^ but is also present in oligodendrocytes and myelin.^13^ In patients with MS, regional NAA levels are reduced in both normal appearing tissue and white matter lesions,^12,14^ making it an important marker of neuronal damage and demyelination. Myo-inositol is primarily synthesized in astrocytes and is used as a marker of glial proliferation and inflammation.^15^ Consequently, regional myo-inositol concentration is increased in white matter lesions and to a lesser extent also in normal appearing tissue in MS.^12^ In addition to metabolic markers of neural and glial metabolism, ^1^H-MRS can also be used to quantify neurotransmitter concentrations of glutamate, the primary excitatory neurotransmitter, and gamma-aminobutyric acid (GABA), the primary inhibitory neurotransmitter in the cortex. ^1^H-MRS of glutamate and GABA has inherent limitations at 3T due to limited spectral resolution. This results in poor sensitivity towards GABA and a lack of specificity for glutamate because of overlapping glutamate and glutamine peaks.^16^ ^1^H-MRS at ultra-high field strength drastically improves the reliability of glutamate and GABA quantification, by increasing signal-to-noise ratio and offering superior spectral resolution.^17^

In this prospective, cross-sectional study, we integrated 7T ^1^H-MRS and high-resolution structural MRI in patients with MS to quantify the regional concentration of glutamate and GABA in the right and left SM1-HAND. We hypothesized that regional cortical involvement, reflected by cortical lesion volume in the SM1-HAND, would shift the regional excitation-inhibition balance in the affected cortical area. We also explored the relationship between regional lesion volume and regional NAA and myo-inositol levels. Here, we hypothesized that, similar to white matter lesions, a larger cortical lesion volume would reduce regional NAA levels in the SM1-HAND voxel, indicating neuronal damage, and increase myo-inositol levels due to gliosis.

## Materials and Methods

### Study participants

We prospectively recruited MS patients with relapsing remitting (RRMS) or secondary progressive (SPMS) MS, aged between 18 and 80 years, from the outpatient clinic at the Danish Multiple Sclerosis Center (Copenhagen University Hospital - Rigshospitalet, Copenhagen, Denmark) between August 2018 and September 2020. Exclusion criteria were clinical relapses, corticosteroid therapy or changes in MS-related medication within 3 months of participation, expanded disability status scale (EDSS) above 7, other neurologic or psychiatric disorders, and contraindications to 7T MRI. Age- and sex-matched healthy volunteers were recruited through advertisements in the same period. All study participants have been described in a previous publication.^11^ The study was approved by the Committees on Health Research Ethics of the Capital Region of Denmark (H-17033372) and monitored by the local good clinical practice unit. Participants gave informed written consent prior to participation. The study complied with the Helsinki declaration of human experimentation and was preregistered at www.clinicaltrials.gov (ID: NCT03653585).

### Experimental design

Data collection was carried out over two experimental sessions within a four-week period. We collected high-resolution structural 7T MRI during the first experimental session and voxel-based 7T ^1^H-MRS during the second. No participants experienced any relapses during their participation.

### Structural MRI and ^1^H-MRS protocol

Structural MRI and ^1^H-MRS were performed on a 7T Achieva scanner (Philips, Best, The Netherlands) using a dual transmit, 32-channel receive head coil (Nova Medical, Wilmington MA, USA). The structural protocol included magnetization prepared fluid attenuated inversion recovery (FLAIR), magnetization prepared rapid gradient echo (MPRAGE), T_2_-weighted turbo spin echo and MP2RAGE. Prospective fat-navigated motion correction^18^ was applied to all structural scans. Detailed MRI parameters can be found in.^11^ ^1^H-MRS measurements were acquired bilaterally from the pericentral “hand knob” regions covering the SM1-HAND.^19^ A cubic 20mm^3^ voxel was manually placed based on a fast T_1_-weighted image (Figure 1). ^1^H-MR spectra were acquired using the semi-localized by adiabatic selective refocusing (sLASER) sequence (TE = 31-33ms, TR = 3175-4960ms, 32 averages, 4000 Hz bandwidth and 2048 data points).^20^ Acquisition details are given in eTable 1.

**Figure 1.**
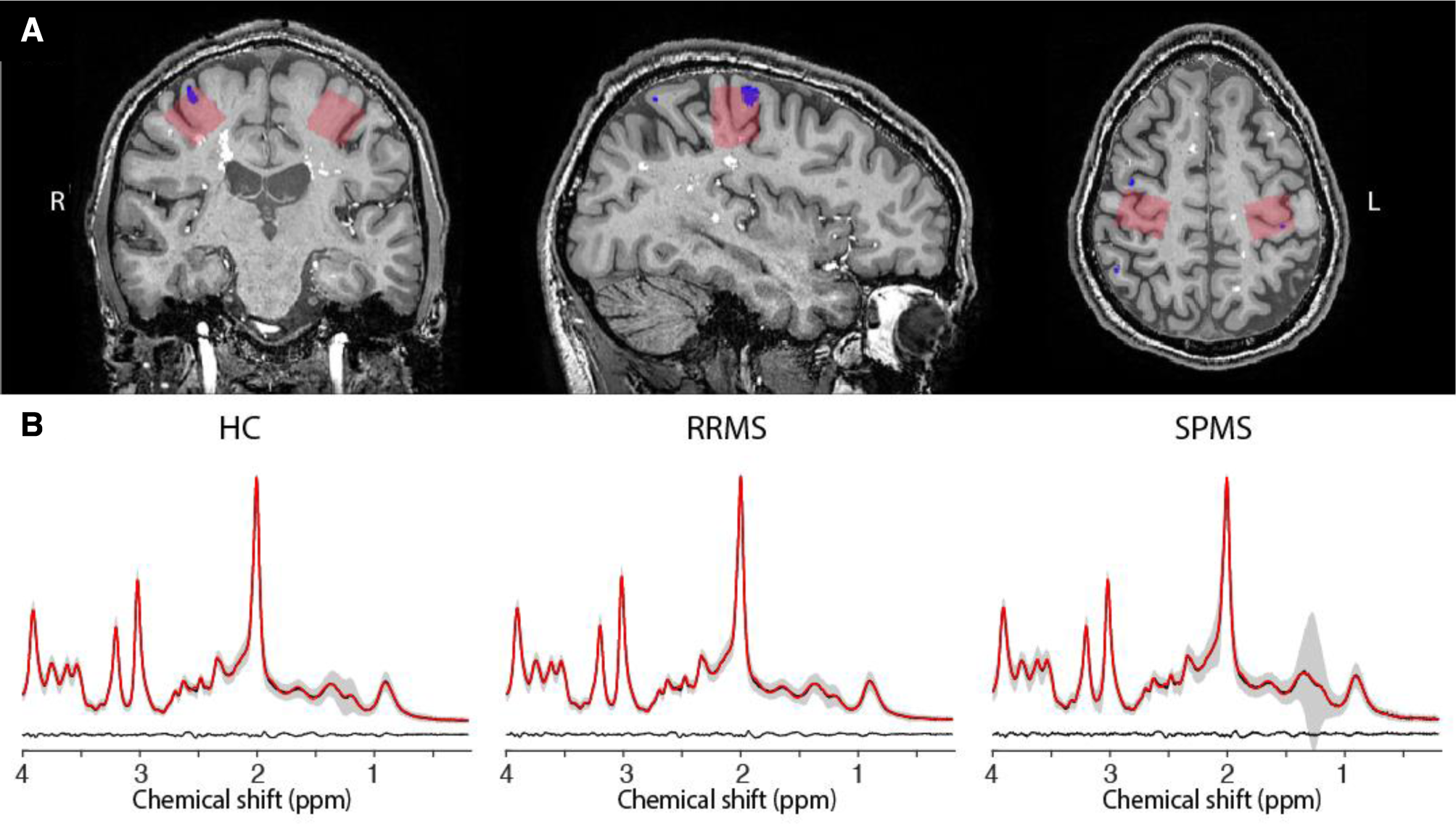
Voxel placement and mean spectra. (**A**) Example MRS voxel placement (red) in the sensorimotor handknob, corresponding to the primary sensorimotor hand area (SM1-HAND). Segmented cortical lesions are shown in blue and white matter lesions in white. (**B**) Average spectra (black) obtained with ^1^H-MRS at 7T. The gray areas representing ± standard deviation for participant groups. Average LCModel fit results (red) and average residuals (black, bottom) are also shown. Abbreviations: HC = healthy controls, RRMS = relapsing remitting multiple sclerosis, SPMS = secondary progressive multiple sclerosis, ppm = parts per million.

### Clinical assessments

We extracted clinical data (EDSS and disease duration) from clinical records, if obtained within three months of study participation. Otherwise, EDSS was assessed on the second experimental session.

#### Processing of structural MRI data

Preprocessing of structural MRI data included bias field correction, resampling to an isotropic voxel size of 0.5 mm^3^, alignment of the brain to MNI space and co-registration as described in.^11^ Cortical- and white matter lesions were manually segmented by three experienced, blinded readers using FLAIR, T_2_-weighted and MPRAGE images as described in.^11^ Whole brain volumetric segmentations were performed with *freesurfer* (version 7.1.1) using MP2RAGE as input image. Lesion filling and manual corrections were performed when needed.

#### Analysis of ^1^H-MRS data

The MR spectra of each voxel were processed using automated in-house Matlab-based (MathWorks, Natick, Mass) software and quantified using LCModel^21^ (version 6.3-1R) over the frequency range from 4.2 ppm to0.2 ppm. The basis set was simulated in FID-A,^22^ modelling 19 metabolites and the macromolecular spectrum (eTable 1). Metabolite concentrations were estimated using the water signal as an internal reference. Tissue correction was applied as described in^23^ with white matter, grey matter, and cerebrospinal fluid tissue fractions calculated from the *freesurfer* segmented images. Water relaxation times were adapted from literature.^24–26^

### Statistical analyses

Differences in demographic- and clinical outcomes were compared with Kruskal-Wallis tests, Mann-Whitney U test or chi-square tests where appropriate. Metabolite concentrations were compared between groups (HC, RRMS, SPMS) using mixed linear models corrected for sex, age and grey- and white matter fraction in the ^1^H-MRS voxel. Subject was used as a random factor with random intercept. Post-hoc multiple comparison corrections were done with the Tukey’s HSD method within each analysis.

The relationship between lesion volumes and metabolite concentrations were assessed in patients only, using mixed linear models with regional glutamate, GABA, NAA, myo-inositol concentrations, patient status (RRMS or SPMS), age and sex as covariates and subject as a random effect with random intercept. Relationships between clinical scores and mean metabolite concentrations of the two hemispheres, were investigated using Spearman’s rank correlations, corrected for multiple comparisons using Tukey’s HSD within each variable. Lesion volumes were log(x+1)-transformed before entering analyses. Statistical analyses were carried out in R-studio (version 2022.07.1) using the packages *lme4, multcomp* and *lmerTest.* Statistical threshold was set at P<0.05 in all analyses.

## Results

### Demographic and clinical characteristics

Thirty-four RRMS and 13 SPMS patients (45.1 +/− 12.5 years, 31 females, median EDSS score: 3.5 [range, 0-6.5], median disease duration: 10 years [range, 0-45]) and 23 healthy controls (44.4 +/− 13 years, 15 female) were studied with 7T MRI. Table 1 summarizes the demographic and clinical variables for each group. MRS data were collected from 94 MS hemispheres and 46 healthy control hemispheres. Nine spectra were discarded because Cramér-Rao lower bounds of GABA were above 50, leaving 88 MS spectra and 43 healthy control spectra. There were no differences in spectral quality metrics or tissue fractions in the ^1^H-MRS voxel between groups (eTable 2). In the patient group, we found a total of 55 cortical lesions in 34 of the 88 SM1-HAND voxels (39%) with a median cortical lesion volume of 9.25 mm^3^ [range, 0.87-203.37 mm^3^]).

**Table 1.** Participant demographics. Abbreviations: HC = healthy control, RRMS = relapsing remitting multiple sclerosis, SPMS = secondary progressive multiple sclerosis, SD = standard deviation, EDSS = expanded disability status scale, FSs = sensory functional systems score, FS = pyramidal functional systems score.

|  | Healthy controls<br>(N=23) | RRMS<br>(N=34) | SPMS<br>(N=13) | All Patients<br>(N=47) |
| --- | --- | --- | --- | --- |
| **Age** |  |  |  |  |
| Mean (SD) | 44.4 (13.0) | 41.9 (11.9) | 53.5 (9.87) | 45.1 (12.5) |
| **Sex** |  |  |  |  |
| Female participants | 15/23 (65.2%) | 23/34 (67.6%) | 8/13 (61.5%) | 31/47 (66.0%) |
| **EDSS score** |  |  |  |  |
| Median [Min, Max] | - | 3.00 [0, 6.50] | 5.00 [3.00, 6.50] | 3.50 [0, 6.50] |
| **FSs** |  |  |  |  |
| Median [Min, Max] | - | 1.00 [0, 3.00] | 2.00 [0, 3.00] | 2.00 [0, 3.00] |
| Missing data | - | 1 (2.9%) | 0 (0%) | 1 (2.1%) |
| **FSp** |  |  |  |  |
| Median [Min, Max] | - | 1.00 [0, 3.00] | 3.00 [2.00, 4.00] | 2.00 [0, 4.00] |
| Missing data | - | 1 (2.9%) | 0 (0%) | 1 (2.1%) |
| **Disease duration** |  |  |  |  |
| Median [Min, Max] | - | 10.0 [0, 35.0] | 17.0 [1.00, 45.0] | 10.0 [0, 45.0] |

### Group differences in metabolite concentrations and clinical correlates

The linear mixed model showed a reduction in regional glutamate concentration in SPMS patients (P=0.009)(figure 2A). Glutamate concentration was not only reduced relative to healthy controls (−0.77 +/− 0.25 mM, P=0.006) but also relative to RRMS patients (−0.66 +/− 0.24 mM, P=0.02). Glutamate concentrations in SM1-HAND correlated negatively with individual EDSS scores (R=-0.39, P=0.03, figure 3A). There were no significant differences in regional GABA concentration between groups (P=0.29)(figure 2B) or significant correlations of mean GABA concentrations with individual EDSS scores in MS patients (R=-28, P=0.17)(figure 3B).

**Figure 2.**
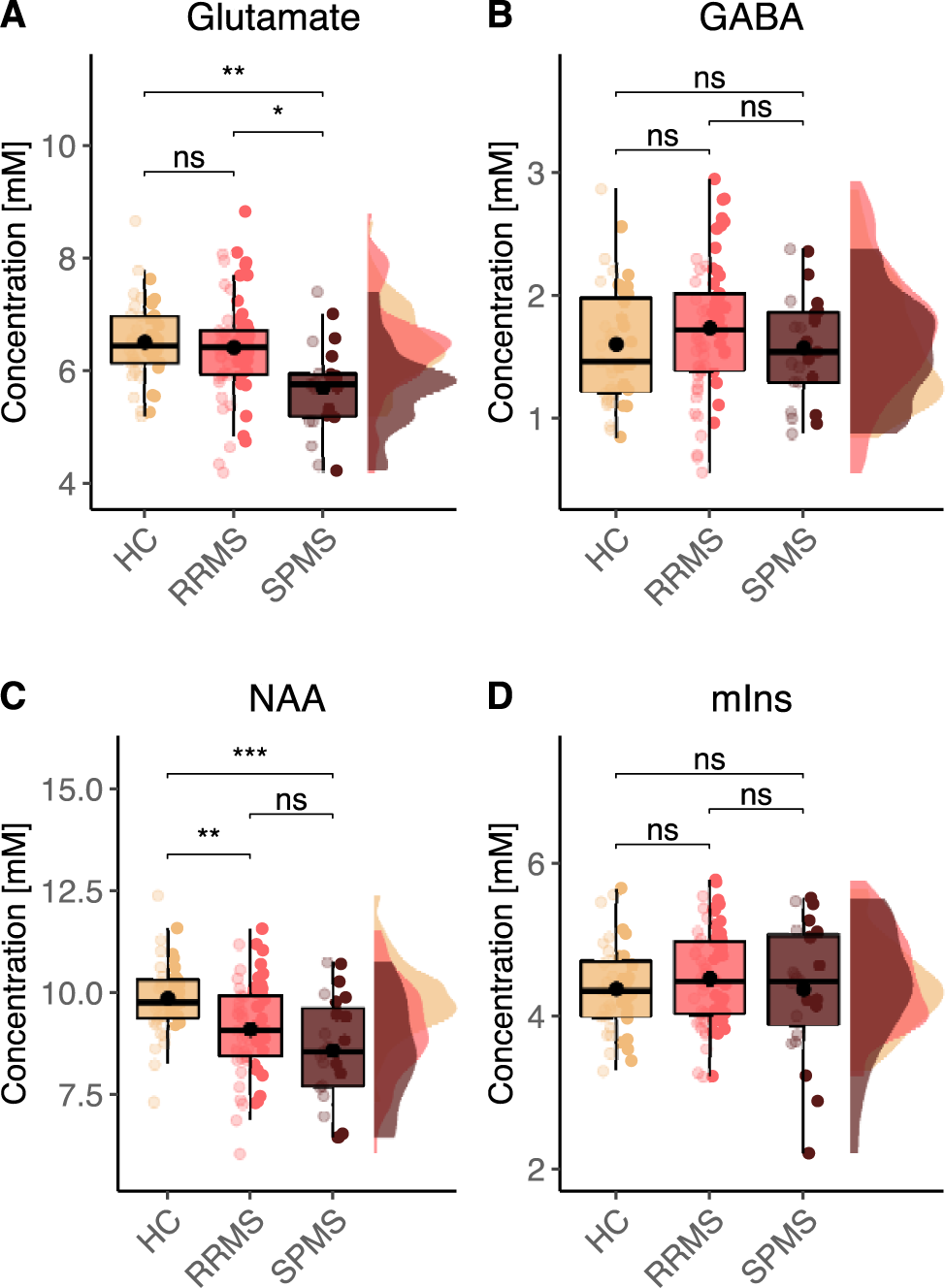
^1^H-MRS metabolite concentrations in SM1-HAND. Box-, point- and density plots of Glutamate (**A**), GABA (**B**), N-acetylaspartate (**C**) and myo-inositol (**D**) between health control participants, RRMS and SPMS patients. Abbreviations: NAA = N-acetylaspartate, mIns = myo-inositol, HC = healthy control, RRMS = relapsing remitting multiple sclerosis, SPMS = secondary progressive multiple sclerosis.

**Figure 3.**
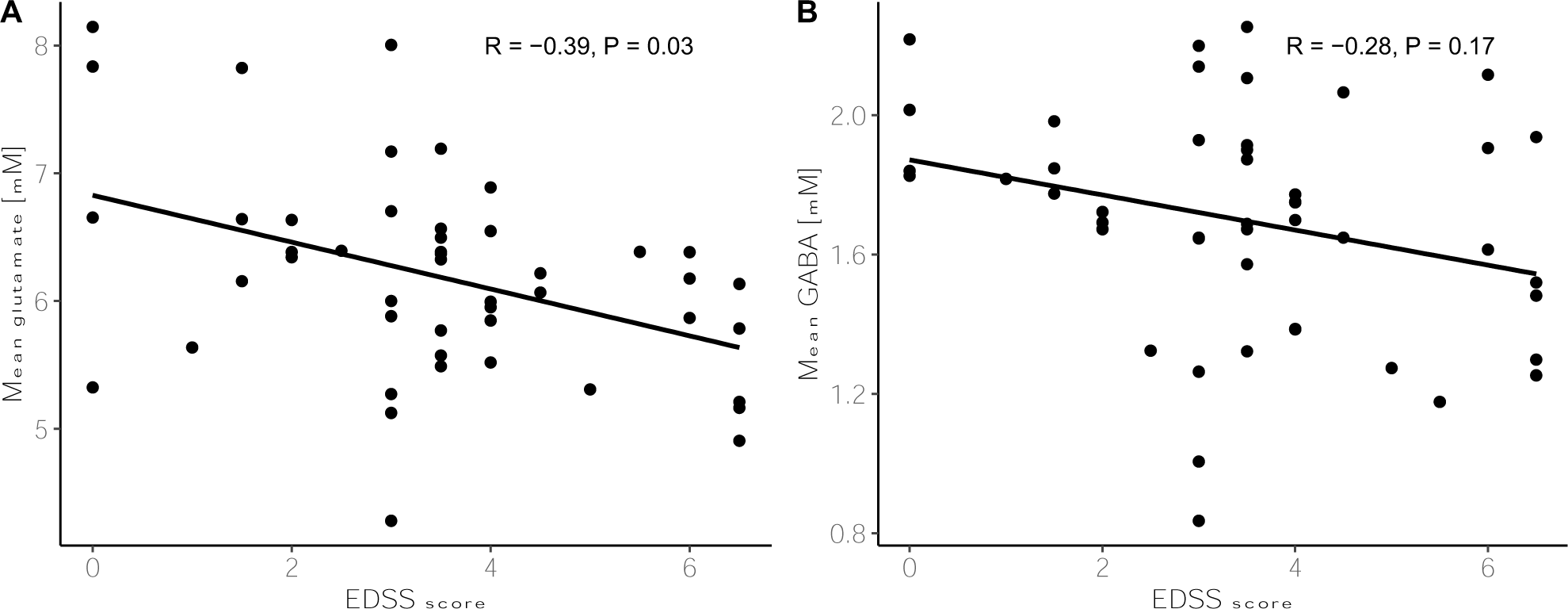
Neurotransmitter concentrations and clinical impairment. Scatterplots showing the correlation between EDSS score (y-axis) and (**A**) mean regional glutamate concentration and (**B**) mean regional GABA concentration from the two MRS voxels of the right and left SM1-HAND voxel measured with ^1^H-MRS at 7T (x-axis).

While regional myo-inositol concentration in SM1-HAND did not differ among groups (P=0.77)(figure 2D), NAA concentrations were lower in MS patients compared to healthy controls (P<0.001)(figure 2C). Mean estimated difference (+/− std. error) was −0.75 +/− 0.24 mM (P=0.005) between RRMS patients and healthy controls and −1.29 +/− 0.33 mM (P<0.001) between SPMS patients and healthy controls. Neither mean NAA concentration (R=-0.18, P=0.47) nor mean myo-inositol concentrations (R=0.01, P=0.93) in SM1-HAND correlated with individual EDSS scores.

### Alterations in metabolite concentrations associated with cortical lesion volume

The linear mixed model showed that cortical lesion volume inside the SM1-HAND was associated with an altered balance between excitatory and inhibitory neurotransmitter concentrations (figure 4A). The higher the regional glutamate concentration, the larger the cortical lesion volume (model estimate +/− std. error, 0.61 +/− 0.21 log(mm^3^), P=0.005). Conversely, the lower the regional GABA concentration, the lower was the cortical lesion volume (−0.71 +/− 0.27 log(mm^3^), P=0.01) in the corresponding MRS voxel. Moreover, regional myo-inositol concentration was associated with higher cortical lesion volume in SM1-HAND (0.48 +/− 0.23 log(mm^3^), P=0.037) and NAA concentration with lower cortical lesion volume (−0.37 +/− 0.15 log(mm^3^), P=0.016)(figure 4A).

**Figure 4.**
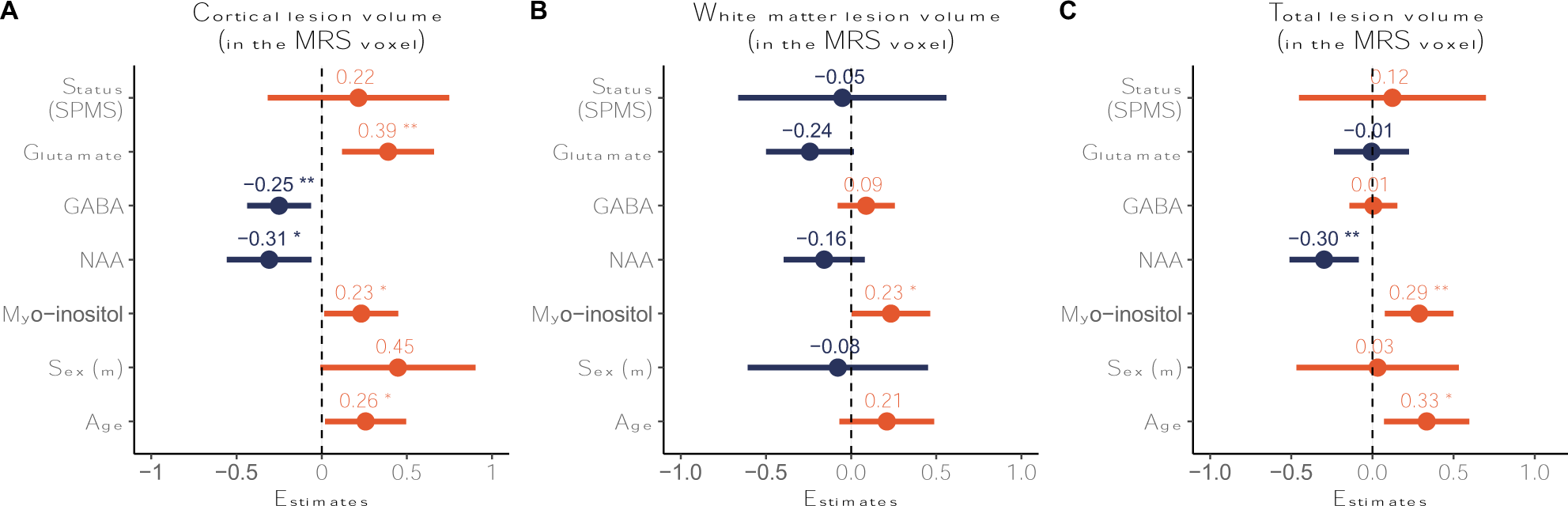
Associations between cortical lesion volume and metabolite concentrations in the ^1^H-MRS voxels covering the right and left SM1-HAND. Standardized beta-coefficients from the mixed linear models with **(A)** cortical lesion volume, **(B)** white matter lesion volume and **(C)** total lesion volume within the ^1^H-MRS voxel as the dependent variable. Abbreviations: MRS = magnetic resonance spectroscopy, SPMS = secondary progressive multiple sclerosis, NAA = N-acetylaspartate.

Because the outcome variable (i.e., regional cortical lesion volume) contained many zeros, we also performed a mixed effects Poisson regression with cortical lesion count in the MRS voxel as the outcome variable and a mixed effects logistic regression with cortical lesion positive/negative MRS voxels in the right or left hemispheres as the outcome variable. These models yielded similar results for regional glutamate (Poisson regression: 0.64 +/− 0.24, P=0.007, logistic regression: 1.46 +/− 0.72, P=0.042) and GABA concentrations (Poisson regression: −0.9 +/− 0.38, P=0.018, logistic regression: −2.15 +/− 0.9, P=0.017), while the effects of regional myo-inositol and NAA concentrations were no longer significant (eTables 3 and 4).

Metabolite concentrations are often referenced to total creatine concentration of the same MRS voxel in order to adjust for variation in spectral quality and partial volume.^12^ This prompted us to perform an additional analysis exploring the relationship between cortical lesion volume and creatine normalized metabolite concentrations. Again, we found similar results for regional glutamate (4.99 +/− 1.43 log(mm^3^), P<0.001) and GABA concentrations (−4.14 +/− 1.58 log(mm^3^), P=0.01). Of the structural MRS markers, only myo-inositol remained statistically significant (4.18 +/− 1.58 log(mm^3^), P=0.01)(eTable 5).

Since the MRS voxel covering right or left SM1-HAND contained both grey and white matter, we computed additional multiple regression models using white matter lesion volume only or the total grey and white matter lesion volume as outcome variables (figure 4B and C). These models showed no significant effects of regional glutamate and GABA concentrations on regional white matter or total lesion volume, indicating that regional cortical lesion fraction rather than regional white matter lesion fraction shifts the balance between excitatory and inhibitory neurotransmitter concentrations. In contrast, the effects of regional NAA and myo-inositol on regional grey matter, white matter or total regional lesion volume were comparable (figure 4B and C) suggesting that both grey and white matter lesions contribute to regional variations in NAA or myo-inositol concentrations in the SM1-HAND voxel.

## Discussion

Using single-voxel 7T ^1^H-MRS of the left and right SM1-HAND, we show that cortical lesion volume is associated with opposite changes in the local concentrations of glutamate and GABA within the affected cortical region, with a regional increase in glutamate and a decrease in GABA concentrations. Additionally, we confirmed and extended previous MR-spectroscopic findings from white matter lesions showing that cortical lesions also associate with MR-spectroscopic metabolic markers of axonal damage (i.e., reduced regional NAA concentration) and astrogliosis (i.e., increased regional myo-inositol concentration).

### Changes in regional glutamate and GABA concentrations

This is, to our knowledge, the first study reporting a relationship between neurotransmitter concentrations and cortical lesions of the same area *in vivo* in patients with multiple sclerosis. Our findings suggest that local cortical lesion load shifts the regional excitation-inhibition balance towards a relative increase in excitation. Histopathological studies have shown that cortical lesions lead to a general loss of cortical neurons compared with normal appearing grey matter.^27^ However, it is less clear whether certain neuronal populations within a cortical lesion are particularly vulnerable. Interestingly, both reductions in GABAergic but not glutamatergic^28^ and glutamatergic, but not GABAergic neuron densities,^29^ have been reported. Similarly, reductions in synapse density specifically related to GABA or glutamate have been shown in human histopathological samples.^30^ It is difficult to relate these *post mortem* measurements of neuron and synapse densities to our *in vivo* measurements of regional neurotransmitter concentrations. Of relevance to our findings, it has been shown that cortical lesions also exhibit reduced glutamate reuptake transporters,^31^ and animal models of MS have shown imbalances between GABA and glutamate concentrations.^32,33^ Together, these studies suggest that the increase in glutamate may be driven by increased glutamate release from activated immune cells, causing glutamate excitotoxicity that in turn leads to oligodendrocyte death and axonal loss.^32,34,35^ Thus, the general reduction of glutamate found in SPMS patients in the current study may reflect general neuropathy of glutamatergic neurons, while local neuroinflammatory processes within cortical lesions or perilesional cortex might have an opposite effect i.e. augmenting glutamate levels. The latter process may be mitigated by restoring cortical levels of GABA which may serve as a compensatory mechanism.^36,37^

A single study has previously combined ^1^H-MRS with lesion measurements at 3T and included grey matter lesion volume as a covariate for predicting GABA concentrations.^38^ In that study, the relationship between regional grey mater lesion volume and GABA concentration was not significant. This null finding suggests that increased sensitivity to detect cortical lesions at 7T may help to improve the clinical utility of cortical lesions in neuroimaging studies on MS.^8^ Another ^1^H-MRS-MRI study at 3T found that higher regional magnetization transfer ratio (MTR) values in the grey- and white matter of the sensorimotor cortex were associated with higher glutamate levels and lower GABA levels.^39^ This relationship between glutamate, GABA and MTR is opposite in sign compared to what would have been expected from the current study. This might not only be due to the lower specificity of 3T MRI but also to the fact that MTR is a rather non-specific microstructural marker associated with changes in regional myelin content but also changes in axons,^40^ and activated microglia.^41,42^

Previous 3T ^1^H-MRS measurements of glutamate and GABA concentrations have yielded conflicting results in MS. Elevated glutamate levels have been shown in normal appearing white matter and acute white matter lesions^43,44^, but reduced levels have also been reported.^39,45,46^ Similarly, regional GABA increases^39^ and decreases^38,47^ have been found in mixed tissue voxels. These discrepancies may be ascribed to glutamate and GABA being difficult to reliably quantify at 3T. Additionally, heterogeneity among studies in terms of acquisition and quantification strategies and clinical characteristics of the experimental cohorts may also have contributed to these discrepant findings.^12^ It is also important to note that these studies have not targeted the motor cortex, but primarily focused on regions within the white matter.

Only one study has so far investigated neurotransmitter concentrations using ultra-high field ^1^H-MRS. In that study, glutamate and GABA levels were reduced in the frontal cortex of patients with progressive MS.^48^ In the current study, we also found an overall reduction in glutamate levels in the SM1-HAND in patients with SPMS, suggesting that regional glutamate concentration decreases after the transition from a relapsing remitting to a progressive course. This finding needs to be corroborated in longitudinal studies.^49^ In contrast, GABA levels in the present study were neither reduced in patients with a relapsing remitting nor a secondary progressive disease course, but cortical lesion volume was found to be associated with lower GABA levels in the SM1-HAND.

### Changes in regional NAA and myo-inositol concentrations

Our combined ^1^H-MRS and lesion measurements at 7T showed a negative relationship between cortical lesion volume and regional NAA concentration, indicating that the level of neuronal damage in the SM1-HAND increases with regional cortical lesion burden. This extends previous ^1^H-MRS studies, which have shown a seemingly progressive reduction in NAA levels from normal appearing regions,^12^ to white matter lesions^50^ and active white matter lesions.^43^ This reduction in NAA levels is most likely related to a reduction in neuronal or axonal density,^51^ but may also reflect local levels of demyelination.^13^

Cortical lesion volume scaled positively with regional myo-inositol concentration, suggesting that regional astrogliosis increases with regional cortical lesion load. Myo-inositol is thought to be an astrocytic metabolic marker^15^ and has commonly been found to be increased in MS, especially in white matter lesions.^12^ This is particularly evident from 7T ^1^H-MRS studies where spectra from an entire brain slab have been acquired simultaneously.^52,53^ Together, these findings confirm previous *in vivo* findings from normal appearing tissue and white matter lesions and histopathological studies showing reduced neuronal density^27^ and astrogliosis within cortical lesions.^54^

### Methodological considerations

The cross-sectional study design makes it difficult to draw any causal conclusions and larger longitudinal studies are warranted to investigate the temporal development of neurotransmitter imbalances from cortical lesions. Another limitation is that metabolite concentrations were measured from a single relatively large voxel with partial volume effects, which are not necessarily comparable between our healthy cohort and MS patients, although there were no differences in tissue fractions between groups.

Even though 7T MRI drastically improves cortical lesion sensitivity, it still only catches around 40%, compared to histopathology.^8^ Thus, despite using state-of-the-art methods, we cannot ascertain that some cortical lesions have not gone unnoticed or that lesion volume has been wrongly estimated.

Previous MRI studies have shown differences in tissue water molarity^55,56^, T_1_ ^57^ and T_2_ ^58^ relaxation times in lesions and normal appearing tissue of MS patients relative to healthy subjects. We believe that these potential differences between groups had minimal impact on our results, because we obtained similar spectroscopic findings with both water and creatine referencing.

## Conclusions

Leveraging the improved sensitivity of ultra-high field MRI for cortical lesion detection and ^1^H-MRS, our study provides novel *in vivo* insight into the metabolic consequences of cortical lesions in multiple sclerosis. We provide evidence that cortical lesions associate with changes in the local balance between glutamate and GABA, as well as changes in markers of neuronal integrity and glial proliferation. Our findings also raise the possibility that glutamate excitotoxicity and reduced intracortical inhibition may be relevant pathophysiological features of cortical lesions in MS. Longitudinal ^1^H-MRS studies and additional neurophysiological measurements of intracortical inhibition and excitation with transcranial magnetic stimulation are warranted to better characterize dynamic changes in excitation-inhibition balance and their link to regional cortical lesion load.

## Data Availability

Pseudonymized data can only be shared with a formal Data Processing Agreement and a formal approval by the Danish Data Protection Agency in line with the requirements of the GDPR.

## Acknowledgements

We would like to thank all the patients and healthy controls who have participated in this project, for their essential support to our research. We would also like to thank Sascha Gude, Helena-Céline Stevelt and Jasmin Merhout for contributing with lesion segmentation, *freesurfer* corrections and quality assurance of MRI data.

## References

1. Reich DS, Lucchinetti CF, Calabresi PA. Multiple Sclerosis. N Engl J Med. 2018;378(2):169–180.

2. Rovira A, Wattjes MP, Tintore M, et al. Evidence-based guidelines: MAGNIMS consensus guidelines on the use of MRI in multiple sclerosis-clinical implementation in the diagnostic process. Nat Rev Neurol. 2015;11(8):471–482.

3. Kutzelnigg A, Lucchinetti CF, Stadelmann C, et al. Cortical demyelination and diffuse white matter injury in multiple sclerosis. Brain. 2005;128(Pt 11):2705–2712.

4. Barkhof F. The clinico-radiological paradox in multiple sclerosis revisited. Curr Opin Neurol. 2002;15(3):239–245.

5. Harrison DM, Roy S, Oh J, et al. Association of Cortical Lesion Burden on 7-T Magnetic Resonance Imaging With Cognition and Disability in Multiple Sclerosis. JAMA Neurol. 2015;72(9):1004–1012.

6. Cocozza S, Cosottini M, Signori A, et al. A clinically feasible 7-Tesla protocol for the identification of cortical lesions in Multiple Sclerosis. Eur Radiol. 2020;30(8):4586–4594.

7. Nielsen AS, Kinkel RP, Madigan N, Tinelli E, Benner T, Mainero C. Contribution of cortical lesion subtypes at 7T MRI to physical and cognitive performance in MS. Neurology. 2013;81(7):641–649.

8. Madsen MAJ, Wiggermann V, Bramow S, Christensen JR, Sellebjerg F, Siebner HR. Imaging cortical multiple sclerosis lesions with ultra-high field MRI. Neuroimage Clin. 2021;32:102847.

9. Kilsdonk ID, Jonkman LE, Klaver R, et al. Increased cortical grey matter lesion detection in multiple sclerosis with 7 T MRI: a post-mortem verification study. Brain. 2016;139(Pt 5):1472–1481.

10. Nielsen AS, Kinkel RP, Tinelli E, Benner T, Cohen-Adad J, Mainero C. Focal cortical lesion detection in multiple sclerosis: 3 Tesla DIR versus 7 Tesla FLASH-T2. J Magn Reson Imaging. 2012;35(3):537–542.

11. Madsen MAJ, Wiggermann V, Marques MFM, et al. Linking lesions in sensorimotor cortex to contralateral hand function in multiple sclerosis: a 7 T MRI study. Brain. 2022.

12. Swanberg KM, Landheer K, Pitt D, Juchem C. Quantifying the Metabolic Signature of Multiple Sclerosis by in vivo Proton Magnetic Resonance Spectroscopy: Current Challenges and Future Outlook in the Translation From Proton Signal to Diagnostic Biomarker. Front Neurol. 2019;10:1173.

13. Nordengen K, Heuser C, Rinholm JE, Matalon R, Gundersen V. Localisation of N-acetylaspartate in oligodendrocytes/myelin. Brain Struct Funct. 2015;220(2):899–917.

14. Caramanos Z, Narayanan S, Arnold DL. 1H-MRS quantification of tNA and tCr in patients with multiple sclerosis: a meta-analytic review. Brain. 2005;128(Pt 11):2483–2506.

15. Ciccarelli O, Barkhof F, Bodini B, et al. Pathogenesis of multiple sclerosis: insights from molecular and metabolic imaging. Lancet Neurol. 2014;13(8):807–822.

16. Tkac I, Andersen P, Adriany G, Merkle H, Ugurbil K, Gruetter R. In vivo 1H NMR spectroscopy of the human brain at 7 T. Magn Reson Med. 2001;46(3):451–456.

17. Prinsen H, de Graaf RA, Mason GF, Pelletier D, Juchem C. Reproducibility measurement of glutathione, GABA, and glutamate: Towards in vivo neurochemical profiling of multiple sclerosis with MR spectroscopy at 7T. J Magn Reson Imaging. 2017;45(1):187–198.

18. Andersen M, Bjorkman-Burtscher IM, Marsman A, Petersen ET, Boer VO. Improvement in diagnostic quality of structural and angiographic MRI of the brain using motion correction with interleaved, volumetric navigators. PLoS One. 2019;14(5):e0217145.

19. Yousry TA, Schmid UD, Alkadhi H, et al. Localization of the motor hand area to a knob on the precentral gyrus. A new landmark. Brain. 1997;120 (Pt 1):141–157.

20. Lin A, Andronesi O, Bogner W, et al. Minimum Reporting Standards for in vivo Magnetic Resonance Spectroscopy (MRSinMRS): Experts’ consensus recommendations. NMR Biomed. 2021;34(5):e4484.

21. Provencher SW. Estimation of metabolite concentrations from localized in vivo proton NMR spectra. Magn Reson Med. 1993;30(6):672–679.

22. Simpson R, Devenyi GA, Jezzard P, Hennessy TJ, Near J. Advanced processing and simulation of MRS data using the FID appliance (FID-A)-An open source, MATLAB-based toolkit. Magn Reson Med. 2017;77(1):23–33.

23. Quadrelli S, Mountford C, Ramadan S. Hitchhiker’s Guide to Voxel Segmentation for Partial Volume Correction of In Vivo Magnetic Resonance Spectroscopy. Magn Reson Insights. 2016;9:1–8.

24. Spijkerman JM, Petersen ET, Hendrikse J, Luijten P, Zwanenburg JJM. T (2) mapping of cerebrospinal fluid: 3 T versus 7 T. MAGMA. 2018;31(3):415–424.

25. Rooney WD, Johnson G, Li X, et al. Magnetic field and tissue dependencies of human brain longitudinal 1H2O relaxation in vivo. Magn Reson Med. 2007;57(2):308–318.

26. Bartha R, Michaeli S, Merkle H, et al. In vivo 1H2O T2+ measurement in the human occipital lobe at 4T and 7T by Carr-Purcell MRI: detection of microscopic susceptibility contrast. Magn Reson Med. 2002;47(4):742–750.

27. Vercellino M, Plano F, Votta B, Mutani R, Giordana MT, Cavalla P. Grey matter pathology in multiple sclerosis. J Neuropathol Exp Neurol. 2005;64(12):1101–1107.

28. Zoupi L, Booker SA, Eigel D, et al. Selective vulnerability of inhibitory networks in multiple sclerosis. Acta Neuropathol. 2021;141(3):415–429.

29. Schirmer L, Velmeshev D, Holmqvist S, et al. Neuronal vulnerability and multilineage diversity in multiple sclerosis. Nature. 2019;573(7772):75–82.

30. Huiskamp M, Kiljan S, Kulik S, et al. Inhibitory synaptic loss drives network changes in multiple sclerosis: An ex vivo to in silico translational study. Mult Scler. 2022;28(13):2010–2019.

31. Vercellino M, Merola A, Piacentino C, et al. Altered glutamate reuptake in relapsing-remitting and secondary progressive multiple sclerosis cortex: correlation with microglia infiltration, demyelination, and neuronal and synaptic damage. J Neuropathol Exp Neurol. 2007;66(8):732–739.

32. Werner P, Pitt D, Raine CS. Multiple sclerosis: altered glutamate homeostasis in lesions correlates with oligodendrocyte and axonal damage. Ann Neurol. 2001;50(2):169–180.

33. Mandolesi G, Gentile A, Musella A, et al. Synaptopathy connects inflammation and neurodegeneration in multiple sclerosis. Nat Rev Neurol. 2015;11(12):711–724.

34. Pitt D, Werner P, Raine CS. Glutamate excitotoxicity in a model of multiple sclerosis. Nat Med. 2000;6(1):67–70.

35. Werner P, Pitt D, Raine CS. Glutamate excitotoxicity--a mechanism for axonal damage and oligodendrocyte death in Multiple Sclerosis? J Neural Transm Suppl. 2000(60):375–385.

36. Gilani AA, Dash RP, Jivrajani MN, Thakur SK, Nivsarkar M. Evaluation of GABAergic Transmission Modulation as a Novel Functional Target for Management of Multiple Sclerosis: Exploring Inhibitory Effect of GABA on Glutamate-Mediated Excitotoxicity. Adv Pharmacol Sci. 2014;2014:632376.

37. Bhat R, Axtell R, Mitra A, et al. Inhibitory role for GABA in autoimmune inflammation. Proc Natl Acad Sci U S A. 2010;107(6):2580–2585.

38. Cawley N, Solanky BS, Muhlert N, et al. Reduced gamma-aminobutyric acid concentration is associated with physical disability in progressive multiple sclerosis. Brain. 2015;138(Pt 9):2584–2595.

39. Nantes JC, Proulx S, Zhong J, et al. GABA and glutamate levels correlate with MTR and clinical disability: Insights from multiple sclerosis. Neuroimage. 2017;157:705–715.

40. Schmierer K, Scaravilli F, Altmann DR, Barker GJ, Miller DH. Magnetization transfer ratio and myelin in postmortem multiple sclerosis brain. Ann Neurol. 2004;56(3):407–415.

41. Schmierer K, Tozer DJ, Scaravilli F, et al. Quantitative magnetization transfer imaging in postmortem multiple sclerosis brain. J Magn Reson Imaging. 2007;26(1):41–51.

42. Moll NM, Rietsch AM, Thomas S, et al. Multiple sclerosis normal-appearing white matter: pathology-imaging correlations. Ann Neurol. 2011;70(5):764–773.

43. Srinivasan R, Sailasuta N, Hurd R, Nelson S, Pelletier D. Evidence of elevated glutamate in multiple sclerosis using magnetic resonance spectroscopy at 3 T. Brain. 2005;128(Pt 5):1016–1025.

44. Azevedo CJ, Kornak J, Chu P, et al. In vivo evidence of glutamate toxicity in multiple sclerosis. Ann Neurol. 2014;76(2):269–278.

45. Muhlert N, Atzori M, De Vita E, et al. Memory in multiple sclerosis is linked to glutamate concentration in grey matter regions. J Neurol Neurosurg Psychiatry. 2014;85(8):833–839.

46. Arm J, Oeltzschner G, Al-Iedani O, Lea R, Lechner-Scott J, Ramadan S. Altered in vivo brain GABA and glutamate levels are associated with multiple sclerosis central fatigue. Eur J Radiol. 2021;137:109610.

47. Kantorova E, Hnilicova P, Bogner W, et al. Neurocognitive performance in relapsing-remitting multiple sclerosis patients is associated with metabolic abnormalities of the thalamus but not the hippocampus-GABA-edited 1H MRS study. Neurol Res. 2022;44(1):57–64.

48. Swanberg KM, Prinsen H, DeStefano K, et al. In vivo evidence of differential frontal cortex metabolic abnormalities in progressive and relapsing-remitting multiple sclerosis. NMR Biomed. 2021;34(11):e4590.

49. MacMillan EL, Tam R, Zhao Y, et al. Progressive multiple sclerosis exhibits decreasing glutamate and glutamine over two years. Mult Scler. 2016;22(1):112–116.

50. Bitsch A, Bruhn H, Vougioukas V, et al. Inflammatory CNS demyelination: histopathologic correlation with in vivo quantitative proton MR spectroscopy. AJNR Am J Neuroradiol. 1999;20(9):1619–1627.

51. Barker PB. Brain Pathology in Multiple Sclerosis with High-Field-Strength MR Spectroscopic Imaging. Radiology. 2022;303(1):151–152.

52. Heckova E, Dal-Bianco A, Strasser B, et al. Extensive Brain Pathologic Alterations Detected with 7.0-T MR Spectroscopic Imaging Associated with Disability in Multiple Sclerosis. Radiology. 2022;303(1):141–150.

53. Donadieu M, Le Fur Y, Lecocq A, et al. Metabolic voxel-based analysis of the complete human brain using fast 3D-MRSI: Proof of concept in multiple sclerosis. J Magn Reson Imaging. 2016;44(2):411–419.

54. Bakshi R, Thompson AJ, Rocca MA, et al. MRI in multiple sclerosis: current status and future prospects. Lancet Neurol. 2008;7(7):615–625.

55. Helms G. Volume correction for edema in single-volume proton MR spectroscopy of contrast-enhancing multiple sclerosis lesions. Magn Reson Med. 2001;46(2):256–263.

56. Sarchielli P, Presciutti O, Tarducci R, et al. Localized (1)H magnetic resonance spectroscopy in mainly cortical gray matter of patients with multiple sclerosis. J Neurol. 2002;249(7):902–910.

57. Sappey-Marinier D. High-resolution NMR spectroscopy of cerebral white matter in multiple sclerosis. Magn Reson Med. 1990;15(2):229–239.

58. West J, Aalto A, Tisell A, et al. Normal appearing and diffusely abnormal white matter in patients with multiple sclerosis assessed with quantitative MR. PLoS One. 2014;9(4):e95161.

