## Supplementary tables for "Linking cortical lesions to metabolic changes in multiple sclerosis using 7T proton MR spectroscopy"

### eTable 1. MRS Acquisition and Post-Processing Parameters

| | |
|---|---|
| **1. Hardware** | |
| a. Field Strength | 7T |
| b. Manufacturer | Philips |
| c. Model | Achieva 7T (R5.1.7 Version C) |
| d. RF Coil | Dual transmit, 32-channel receive head coil (Nova Medical, Wilmington MA, USA) |
| **2. Acquisition** | |
| a. Pulse Sequence | sLASER with FOCI refocusing |
| b. VOI Location | Sensorimotor cortex |
| c. Nominal Voxel Size | 2x2x2 cm3 |
| d. TR | 3175ms (81)/3700ms (28)/4960ms (12) |
| e. TE | 31-33 ms |
| f. Number of Averages | 32 |
| g. Spectral Width/Data Points | 4kHz/2048 |
| h. Frequency Offset | 3 ppm |
| i. Water Suppression | VAPOR |
| j. Shimming | Second-order shimming was carried out using a field map–based shimming algorithm ("shimtool") |
| k. Triggering | no |
| **3. Data Analysis** | |
| a. Analysis Software | LCModel |
| b. Processing Steps | Coil Combination, Phase Correction, Averaging |
| c. Output Measure | Tissue-corrected water reference / creatine reference |
| d. Model fitting | Basis set with 19 metabolites and measured MM* |
| d.1 Metabolites | Alanine, Ascorbate, Aspartate, Creatine, GABA, Glucose, Glutamine, Glutamate, Glycerophosphocholine, Glutathione, Myo-inositol, Lactate, N-acetylaspartate, N-acetylaspartylglutamate, Phosphocholine, Phosphocreatine, Phosphoetanolamine, Scyllo-inositol, Taurine |
| **4. Data Quality** | |
| a. Reported QA Variables | SNR, linewidth and CRLB as reported from LCModel |
| b. Data Exclusion Criteria | CRLBs of the reported metabolites above 50% |
| c. QA of Postprocessing | See supplementary table 2 |
| d. Sample Spectrum | See figure 1B |

| | Healthy Controls (N=43) | RRMS (N=65) | SPMS (N=23) | All Patients (N=88) |
|---|---|---|---|---|
| **FWHM (Hz)** | | | | |
| Mean (SD) | 4.32 (0.823) | 4.36 (1.09) | 4.64 (1.51) | 4.43 (1.21) |
| **SNR** | | | | |
| Mean (SD) | 44.4 (7.77) | 41.0 (8.33) | 41.3 (10.2) | 41.1 (8.78) |
| **CRLB % (GABA)** | | | | |
| Median [Min, Max] | 22.0 [10.0, 35.0] | 17.0 [9.00, 50.0] | 20.0 [11.0, 33.0] | 17.5 [9.00, 50.0] |
| **CRLB % (Glutamate)** | | | | |
| Median [Min, Max] | 3.00 [2.00, 5.00] | 3.00 [2.00, 5.00] | 3.00 [3.00, 5.00] | 3.00 [2.00, 5.00] |
| **CRLB % (NAA)** | | | | |
| Median [Min, Max] | 2.00 [1.00, 3.00] | 2.00 [1.00, 3.00] | 2.00 [1.00, 4.00] | 2.00 [1.00, 4.00] |
| **CRLB % (Myo-inositol)** | | | | |
| Median [Min, Max] | 4.00 [3.00, 8.00] | 4.00 [3.00, 7.00] | 4.00 [3.00, 9.00] | 4.00 [3.00, 9.00] |
| **Grey matter fraction (%)** | | | | |
| Mean (SD) | 32.9 (3.81) | 33.1 (3.80) | 30.8 (4.48) | 32.5 (4.09) |
| **White matter fraction (%)** | | | | |
| Mean (SD) | 54.9 (5.84) | 52.4 (5.96) | 54.8 (7.04) | 53.0 (6.31) |

**eTable 2. Magnetic resonance spectroscopy quality metrics.** Abbreviations: HC = healthy controls, RRMS = relapsing remitting multiple sclerosis, SPMS = secondary progressive multiple sclerosis, FWHM = Full width at half max, SNR = signal to noise ratio, CRLB = Cramer-Rao lower bounds, NAA = N-acetylaspartate.

| **CL+/- within the MRS voxel** | | | |
|---|---|---|---|
| *Predictors* | *Odds Ratios* | *CI* | *p* |
| (Intercept) | 0.00 | 0.00 – 0.95 | **0.048** |
| SPMS | 1.62 | 0.23 – 11.52 | 0.629 |
| **Glutamate** | **4.31** | **1.06 – 17.59** | **0.042** |
| **GABA** | **0.12** | **0.02 – 0.68** | **0.017** |
| NAA | 0.52 | 0.22 – 1.26 | 0.148 |
| Myo-inositol | 1.70 | 0.54 – 5.39 | 0.366 |
| **Sex [male]** | **1.10** | **1.01 – 1.20** | **0.030** |
| Age | 2.99 | 0.54 – 16.66 | 0.211 |
| **Random Effects** | | | |
| $\sigma^2$ | 3.29 | | |
| $\tau_{00\ subject}$ | 2.17 | | |
| ICC | 0.40 | | |
| $N_{subject}$ | 47 | | |
| Observations | 88 | | |
| Marginal $R^2$ / Conditional $R^2$ | 0.343 / 0.605 | | |

**eTable 3. Model summary of the mixed effects logistic regression model on CL+/- group data.** Abbreviations: CL = cortical lesion, MRS = magnetic resonance spectroscopy, CI = confidence interval, SPMS = secondary progressive multiple sclerosis, ICC = intra class correlations, NAA = N-acetylaspartate.

| **Cortical lesion number within the MRS voxel** | | | |
|---|---|---|---|
| *Predictors* | *Incidence Rate Ratios* | *CI* | *p* |
| (Intercept) | 0.00 | 0.00 – 0.09 | **<0.001** |
| SPMS | 1.02 | 0.48 – 2.19 | 0.956 |
| **Glutamate** | **1.90** | **1.18 – 3.06** | **0.008** |
| **GABA** | **0.41** | **0.19 – 0.87** | **0.020** |
| NAA | 0.78 | 0.55 – 1.11 | 0.168 |
| Myo-inositol | 1.48 | 0.89 – 2.46 | 0.126 |
| Sex [M] | 1.79 | 0.90 – 3.55 | 0.096 |
| **Age** | **1.05** | **1.02 – 1.09** | **0.001** |
| **Random Effects** | | | |
| $\sigma^2$ | 1.21 | | |
| $\tau_{00\ subject}$ | 0.15 | | |
| ICC | 0.11 | | |
| $N_{subject}$ | 47 | | |
| Observations | 88 | | |
| Marginal $R^2$ / Conditional $R^2$ | 0.333 / 0.408 | | |

**eTable 4. Model summary of the mixed effects Poisson regression model on cortical lesion count data.** Abbreviations: MRS = magnetic resonance spectroscopy, CI = confidence interval, SPMS = secondary progressive multiple sclerosis, ICC = intra class correlations, NAA = N-acetylaspartate.

| **Cortical lesion volume within the MRS voxel (log+1 transformed)** | | | |
|---|---|---|---|
| *Predictors* | *Estimates* | *CI* | *p* |
| (Intercept) | -5.97 | -11.60 – -0.34 | **0.038** |
| SPMS | 0.13 | -0.63 – 0.89 | 0.738 |
| **Glutamate/tCr** | 4.99 | 2.15 – 7.83 | **0.001** |
| **GABA/tCr** | -4.14 | -7.28 – -1.00 | **0.011** |
| NAA/tCr | -1.45 | -3.68 – 0.78 | 0.199 |
| **Myo-inositol/tCr** | 4.18 | 1.04 – 7.33 | **0.010** |
| **Sex [male]** | 0.74 | 0.08 – 1.41 | **0.028** |
| **Age** | 0.04 | 0.01 – 0.07 | **0.008** |
| **Random Effects** | | | |
| $\sigma^2$ | 0.97 | | |
| $\tau_{00\ subject}$ | 0.49 | | |
| ICC | 0.34 | | |
| $N_{subject}$ | 47 | | |
| Observations | 88 | | |
| Marginal $R^2$ / Conditional $R^2$ | 0.286 / 0.526 | | |

**eTable 5. Model summary of the linear mixed effects model on cortical lesion volume with creatine normalized metabolite concentrations.** Abbreviations: MRS = magnetic resonance spectroscopy, CI = confidence interval, SPMS = secondary progressive multiple sclerosis, tCr = total creatine, ICC = intra class correlations, NAA = N-acetylaspartate.
